# Puberty Timing and Markers of Cardiovascular Structure and Function at 25 Years: A Prospective Cohort Study

**DOI:** 10.1101/2020.11.25.20238493

**Authors:** Gillian M. Maher, Lisa Ryan, Fergus P. McCarthy, Alun Hughes, Chloe Park, Abigail Fraser, Laura D. Howe, Patricia M. Kearney, Linda M. O’Keeffe

**Affiliations:** INFANT Research Centre, University College Cork, Cork, Ireland; School of Public Health, University College Cork, Cork, Ireland; Department of Obstetrics and Gynaecology, University College Cork, Cork, Ireland; Department of Population Science and Experimental Medicine, Institute of Cardiovascular Science, Faculty of Population Health Sciences, University College London, Gower Street, London WC1E 6BT, United Kingdom; MRC Unit for Lifelong Health and Ageing at University College London, London, United Kingdom; Population Health Sciences, Bristol Medical School, Oakfield House, Oakfield Grove, Bristol, UK, BS82BN

**Keywords:** Puberty, Cardiovascular Structure, ALSPAC

## Abstract

**Importance:** Whether earlier onset of puberty is associated with higher cardiovascular risk in early adulthood is not well understood.

**Objective:** To examine the association between puberty timing and markers of cardiovascular structure and function at age 25 years.

**Design:** Prospective birth cohort study.

**Setting:** The Southwest region of England.

**Participants:** Participants in the Avon Longitudinal Study of Parents and Children (ALSPAC), born between April 1, 1991, and December 31, 1992.

**Exposure:** Age at peak height velocity (aPHV), an objective and validated growth-based measure of puberty onset.

**Main Outcomes and Measures:** Cardiovascular structure and function at age 25 years: carotid intima-media thickness (CIMT), left ventricular mass index (LVMI) and relative wall thickness (RWT), pulse wave velocity (PWV) and systolic blood pressure (SBP). Multiple imputation was used to impute missing data on covariates and outcomes. Linear regression was used to examine the association between aPHV and each measure of cardiac structure and function, adjusting for maternal age, gestational age, household social class, maternal education, mother’s partner’s education, breastfeeding, parity, birthweight, maternal body mass index, maternal marital status, maternal prenatal smoking status, and height and fat mass at age 9. All analyses were stratified by sex.

**Results:** A total of 2752-4571 participants were included in the imputed analyses. A one-year older aPHV was not strongly associated with markers of cardiac structure and function in males and females at 25 years and most results spanned the null value. In adjusted analyses a one-year older aPHV was associated with 0.003mm (95% Confidence Interval (CI): 0.00001, 0.006) and 0.0008mm (95% CI: −0.002, 0.003) higher CIMT; 0.02m/s (95% CI: −0.05, 0.09) and 0.02m/s (95% CI: −0.04, 0.09) higher PWV; and 0.003mmHg (95% CI: −0.60, 0.60) and 0.13mmHg (95% CI: −0.44, 0.70) higher SBP, among males and females respectively. A one-year older aPHV was associated with −0.55g/m^2.7^ (95% CI: −0.03, −1.08) and −0.89g/m^2.7^ (95% CI: −0.45, −1.34) lower LVMI and −0.001 (95% CI: −0.006, 0.002) and −0.002 (95% CI: −0.006, 0.002) lower RWT among males and females.

**Conclusions and Relevance:** Earlier puberty is unlikely to have a major impact on pre-clinical cardiovascular risk in early adulthood.

**Key Points:** *Question:* Is puberty timing associated with cardiovascular structure and function at age 25 years?

*Findings:* Adjusted estimates from this prospective birth cohort study suggest no strong evidence of association between age at puberty (measured using age at peak height velocity) and carotid intima-media thickness (CIMT), left ventricular mass index (LVMI), and relative wall thickness (RWT), pulse wave velocity (PWV), and systolic blood pressure (SBP) at age 25 years among males and females, with results spanning the null in all but LVMI.

*Meaning:* Earlier puberty is unlikely to have a major impact on pre-clinical cardiovascular risk in early adulthood.

## Introduction

Cardiovascular disease (CVD) is a major cause of morbidity and mortality worldwide, with 7.8 million premature CVD deaths estimated in 2025 if current trajectories are not altered^(1, 2)^. CVD risk originates in early life and tracks through the life course^(3, 4)^. Onset of puberty is a transitional period between childhood and adulthood resulting in physical bodily changes including rapid growth, and intense hormonal activity^(5, 6)^. Age at puberty onset has been decreasing for several decades, with increasing childhood adiposity (a condition of being severely overweight, or obese) thought to play a substantial role^(7)^.

Several studies to date have examined the association between earlier puberty timing and CVD risk, with conflicting findings^(8-14)^. A key limitation of many studies including observational cohort studies and Mendelian Randomisation (MR) designs has included lack of adjustment for early childhood adiposity or use of indirect measures of adiposity such as BMI for adjustment, resulting in residual confounding of the puberty timing-CVD risk associations by early life adiposity^(8, 9, 11, 12, 15, 16)^. In addition, though age at menarche offers a reliable marker of puberty timing in females, most previous studies have assessed puberty timing using self-reported measures among males such as voice change, facial hair and pubic hair^(17, 18)^, leading to measurement error^(19)^. Thus, studies examining the association between objectively measured puberty timing and CVD risk in males and females, while adjusting for direct measures of childhood adiposity are required to better understand the aetiology of puberty timing and CVD risk in early life.

Using an objective growth-based measure of puberty onset (age at peak height velocity [aPHV]), we aimed to better understand the association between puberty timing and pre-clinical cardiovascular risk in early adulthood using markers of cardiovascular structure and function at age 25 years (carotid intima-media thickness [CIMT], left ventricular mass index [LVMI] and relative wall thickness [RWT], pulse wave velocity [PWV] and systolic blood pressure [SBP]).

## Methods

### Study Participants

We used data from the Avon Longitudinal Study of Parents and Children (ALSPAC), a prospective birth cohort study based in Southwest England^(20-22)^. The women invited to participate in this study were pregnant with an expected delivery date between April 1, 1991, and December 31, 1992, and living in one of the three Bristol-based health districts. A detailed description of this study is available elsewhere^(20-22)^.

The initial number of pregnancies enrolled was 14,541. When the oldest children were approximately 7 years of age, an attempt was made to increase the initial sample with eligible children who did not join the study originally. This resulted in an additional 913 children being enrolled. Therefore, the total sample size was 15,454 pregnancies. Of these 14,901 were alive at 1 year of age. In the three decades since enrolment, ALSPAC has used questionnaires completed by both parents and children, routine medical data and research clinics as methods of follow-up. The clinics took place when the participants were 7, 9, 10, 11, 13, 15, 17 and 25 years old^(20-24)^.

Ethical approval for the ALSPAC study was obtained from the ALSPAC Law and Ethics Committee and Local Research Ethics Committees. Informed consent for the use of data collected via questionnaires and clinics was obtained from participants following the recommendations of the ALSPAC Law and Ethics Committee at the time. The study website contains details of all the data that is available through a fully searchable data dictionary and variable search tool: http://www.bristol.ac.uk/alspac/researchers/our-data/^(25)^.

### Exposure

#### Assessment of Timing of Puberty

Puberty is a period of intense hormonal activity and rapid growth, of which the most striking feature is the spurt in height^(6)^. aPHV is a validated measure of puberty timing^(6)^ captured using Superimposition by Translation and Rotation (SITAR), a non-linear multilevel model with natural cubic splines which estimates the population average growth curve and departures from it as random effects^(26, 27)^. Using SITAR, PHV was identified in ALSPAC participants using numerical differentiation of the individually predicted growth curves, with aPHV being the age at which the maximum velocity is observed^(26-28)^. Repeated height data included measurements from research clinics and were used here to derive aPHV using SITAR. Individuals with at least one measurement of height from 5 years to <10 years, 10 years to < 15 years and 15years to 20 years were included. Data were analysed for males and females separately. The model was fitted using the SITAR package in R version 3.4.1, as described elsewhere^(28)^. Further details on how aPHV was derived is described elsewhere^(28, 29)^.

### Outcomes

#### Assessment of Cardiovascular Risk

Carotid intima-media thickness (CIMT), left ventricular mass index (LVM) and relative wall thickness (RWT), pulse wave velocity (PWV), and systolic blood pressure (SBP) were measured at research clinics at age 25 years. Participants fasted for six hours before the clinic, with the exception of those participants with a diagnosis of diabetes or a condition that would not allow fasting. CIMT scans of the left and right common carotid arteries were performed using a CardioHealth Panasonic system with a 13-5 MHz linear array broadband transducer according to a standardised protocol. CIMT has been validated in several studies as a strong predictor of CVD-risk^(30-32)^. Participants lay on a couch with their arms by their side, while a trained researcher performed the ultrasound test on both sides of their neck. Right and left CIMT measurements were taken to be the average of 3 end-diastolic measurements of the far-wall of the common carotid artery over a length of 5-10mm, and 10□mm adjoining the bifurcation. The mean of both right and left CIMT measures was calculated and used here.

Echocardiography was performed by two experienced echo-cardiographers using a Philips EPIQ 7G Ultrasound System equipped with a X5-1 transducer in accordance with American Society of Echocardiography guidelines; these techniques are described elsewhere^(33)^. Left ventricular mass (LVM), assessed by ultrasound, was indexed to height^2.7^ to adjust for body surface area. RWT was calculated using left ventricular internal diameter in diastole and thickness of the left ventricular posterior wall and septal wall. PWV was measured using a Vicorder device (Skidmore Medical, Bristol, UK) at femoral and carotid artery level which has been validated in previous studies in adolescents^(34)^. Ten PWV measurements were taken within ≤0.5□m/s of each other. These were averaged to give a measurement of arterial stiffness. Systolic blood pressure was measured (typically from the right arm) with the subject in a sitting position using an Omron 705-IT machine^(33)^.

### Covariates

We considered the following as possible confounders of the association between aPHV and cardiovascular structure and function at age 25 years; maternal age, gestational age at birth, household social class, maternal education, mother’s partner’s education, breastfeeding of baby until three months, parity, birthweight, maternal body mass index (BMI), maternal marital status, maternal smoking status during first 3 months of pregnancy, and height and DXA-determined fat mass at age 9. See eMethods 1 in the Supplement for description and measurement of confounders.

### Statistical Analysis

Statistical analysis was performed using Stata MP 14.2. Linearity of association between aPHV and cardiovascular structure and function in males and females was assessed by comparing fit of models regressing CIMT, LVMI, RWT, PWV and SBP on fourths of aPHV (treated as a continuous exposure) to models regressing CIMT, LVMI, RWT, PWV and SBP on fourths of aPHV (treated as a categorical exposure); models were then formally compared using a likelihood ratio test. Data on aPHV and all cardiovascular outcomes were normally distributed. We used linear regression to examine the association between aPHV and each measure of cardiac structure and function. All analyses were stratified by sex to examine whether associations of aPHV and each outcome differed for males and females.

#### Dealing with Missing Data

There were 1197-2193 participants with complete data for aPHV, each outcome and all covariates. Cardiac structure and function were also measured at the 18 year ALSPAC clinic. To increase efficiency and minimise selection bias, we used multivariate multiple imputation to impute missing data on covariates and outcomes in all participants that had a measure of aPHV and had a measure of the outcome at the age 18 year or age 25 year research clinic (N=4339 for CIMT, N=2752 for LVMI, N=2776 for RWT, N=3964 for PWV and N=4571 for SBP). We carried out 20 cycles of regression switching and generated 20 imputation datasets^(35)^. We then examined associations of aPHV and outcomes in these multiple imputed datasets; results are averaged across the results from each of these 20 datasets using Rubin’s rules, taking account of uncertainty in the imputation so that the standard errors for any regression coefficients (used to calculate 95% confidence intervals) take account of uncertainty in the imputations and uncertainty in the estimate^(35)^. See eTable 1 for a list of the variables included in the multiple imputation models and how they were entered into the models. We repeated the main adjusted analysis in the observed dataset, among participants with complete-case data on exposure, outcome and covariates (N=1199 for CIMT, N=1197 for LVMI, N=1203 for RWT, N=1394 for PWV and N=2193 for SBP).

**Table 1:**
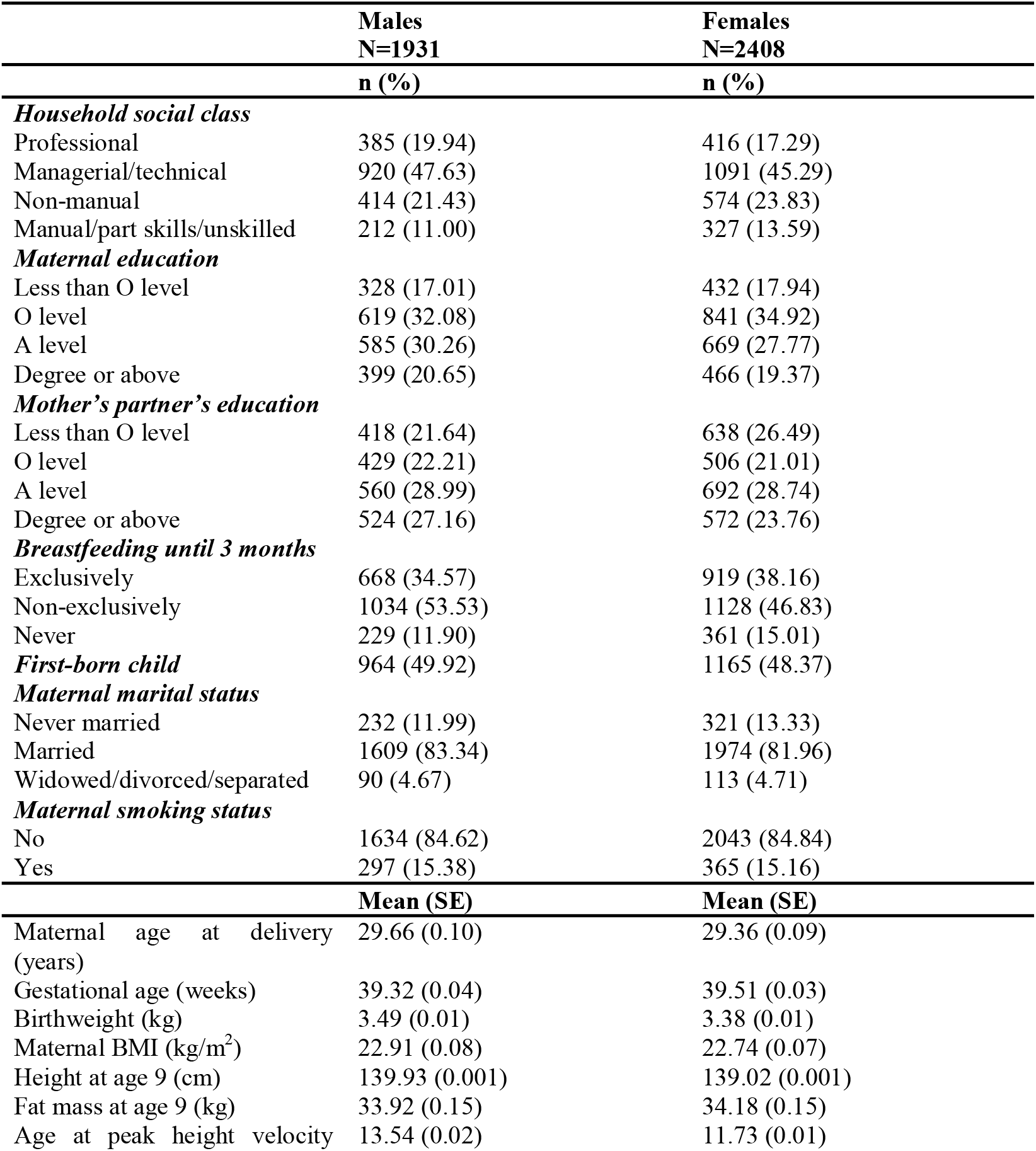
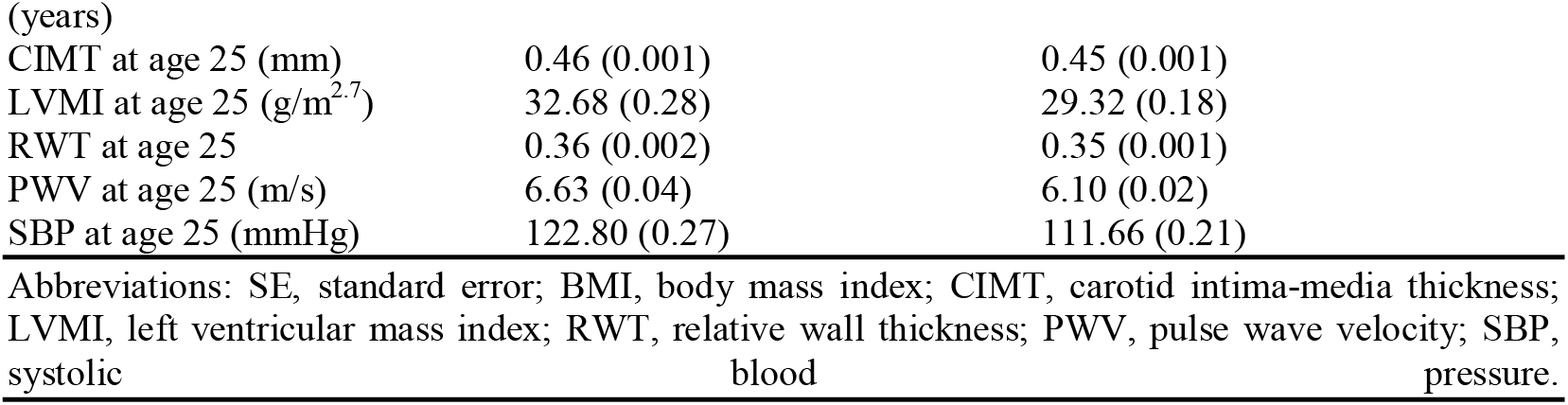
Characteristics of ALSPAC participants included in the analysis by sex and based on imputed data.

## Results

The characteristics of the cohort are shown in Table 1. A total of 2752-4571 participants were included in the imputed analyses. The mean aPHV was 13.54 years in males and 11.73 years in females. Findings from linearity tests of the association between aPHV and cardiac structure and function demonstrated little evidence of departures from linearity, allowing aPHV to be examined as a continuous exposure (eTable 2). The distribution of variables in observed and imputed data is shown in eTables 3-7; distributions of most variables were broadly similar between observed and imputed datasets with some minor differences only for socio-economic position indicators which were slightly higher in the observed datasets.

**Table 2:**
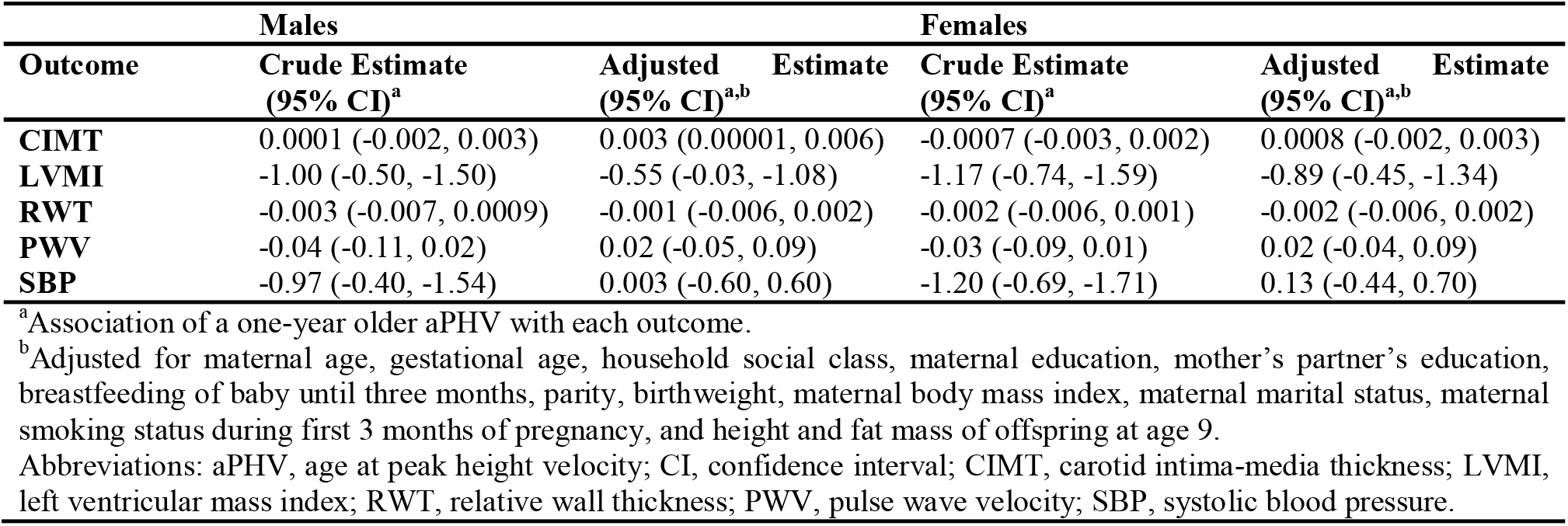
Associations between age at peak height velocity and cardiac structure and function in males and females at age 25.

### Age at Peak Height Velocity and Cardiovascular Structure and Function

#### Males

There was little evidence of association between aPHV and cardiac structure and function in males, with results mostly spanning the null value. For example, in confounder-adjusted analyses, a one-year older aPHV was associated with 0.003mm (95% Confidence Interval (CI): 0.00001, 0.006) higher CIMT, 0.02m/s (95% CI: −0.05, 0.09) higher PWV and 0.003mmHg (95% CI: −0.60, 0.60) higher SBP at 25 years. A one-year older aPHV was associated with a −0.55g/m^2.7^ (95% CI: −0.03, −1.08) lower LVMI and −0.001 (95% CI: - 0.006, 0.002) lower RWT at 25 years.

### Females

Similar to males, there was little evidence of association observed between aPHV and cardiac structure and function in females, with results mostly spanning the null value. In confounder-adjusted analyses, a one-year older aPHV was associated with 0.0008mm (95% Confidence Interval (CI): −0.002, 0.003) higher CIMT, 0.02m/s (95% CI: −0.04, 0.09) higher PWV and 0.13mmHg (95% CI: −0.44, 0.70) higher SBP at 25 years. A one-year older aPHV was associated with a −0.89g/m^2.7^ (95% CI: −0.45, −1.34) lower LVMI and −0.002 (95% CI: - 0.006, 0.002) lower RWT at 25 years.

Unadjusted and confounder adjusted results were not materially different for each outcome in males and females (Table 1).

Adjusted associations of aPHV with measures of cardiac structure and function among participants with complete-case data on exposure, outcome and covariates were comparable to the main results obtained using imputed datasets (eTable 8).

## Discussion

This study aimed to better understand the association between puberty timing and cardiovascular structure and function at age 25 years using aPHV as an objective growth-based measure of puberty onset and CIMT, LVMI, RWT, PWV and SBP as pre-clinical measures of cardiovascular risk. There was little evidence of association between aPHV and cardiovascular structure and function at 25 years, with results spanning the null in all but LVMI for which the association was inverse.

Previous studies have shown that a ∼ 5g/m^2^ higher LVMI can be predicted to correspond to a 7–20% increase in CVD morbidity and mortality^(36-38)^. Given the small differences in LVMI per year older aPHV found here, our findings suggest that earlier puberty is unlikely to have a major impact on pre-clinical cardiovascular risk in early adulthood.

### Comparison with other studies

Several studies to date have examined the association between puberty timing and clinical endpoints such as CVD and traditional cardiovascular risk factors such as blood pressure and adiposity, with conflicting findings^(8-10, 15, 17, 18, 39)^. However, these studies used different measures of puberty timing (objective vs. self-report)^(39, 40)^, had varying degrees of adjustment for confounding (particularly early childhood adiposity)^(8-10, 15)^, and different ranges in follow-up and age of measurement of outcomes^(14, 18, 41)^.

In contrast, fewer studies have examined associations of puberty timing with measures of cardiac structure and function, which are used clinically to assess arterial stiffness, left ventricular hypertrophy and cardiac remodelling^(40)^. Hardy and colleagues found evidence of an association between later puberty timing (measured using age at menarche) and lower LVM and RWT in females in the National Study of Health and Development (NSHD) (N=1,385), though results attenuated after adjustment for childhood or adult adiposity^(40)^. In contrast, there was no strong evidence of an association between puberty timing (measured via physical examination) and any measure of cardiac structure or function in males in the NSHD, a finding comparable to ours^(40)^. Our findings are also comparable to results from the Young Finns Cohort (N=794) which demonstrated no strong evidence of associations between puberty timing (measured via age at menarche) and CIMT in females at age 30-39 years^(14)^. Similarly, in a study of 800 women aged 50 to 81 years in Southern Germany, age at menarche was not associated with CIMT in both unadjusted and confounder-adjusted analyses^(41)^.

### Strengths and Limitations

Strengths of this study include use of an objective measure of puberty timing (aPHV) based on prospective, repeated measures of height from age 5 years to 20 years which is a more accurate marker of puberty timing than self-reported measures. We also used measures of pre-clinical CVD, which strongly predict later cardiovascular risk. We adjusted for childhood adiposity using DXA fat mass at age 9, allowing us to adjust for directly measured pre-pubertal adiposity, a key limitation in several previous studies^(8-10, 15)^. However, there are also several limitations; a limitation of our adiposity adjustment may include the possibility that DXA fat mass is measured after pubertal onset for a small proportion of participants (N=1 for males, N=41 for females) giving rise to the possibility of collider bias if DXA fat mass at age 9 is a mediator of the puberty timing-cardiac risk association. However, we believe that any bias introduced due to this is minimal, given that prior work in this cohort has shown that puberty has little impact on post-pubertal fat mass gain^(29)^ (suggesting fat mass is an unlikely mediator of any associations of puberty timing with health outcomes) and that analyses without adjustment for fat mass were similar to confounder-adjusted results. Further limitations include potential for selection bias due to missing data and attrition from the cohort. However, the use of multivariate multiple imputation aimed to minimise bias by imputing missing covariate and outcome data, and results in multiply imputed and complete-case data were similar. Finally, the vast majority of the ALSPAC cohort are of White ethnicity^(20)^, and a key limitation of our study is the generalisability of the findings to non-White ethnicities.

## Conclusion

Age at peak height velocity was not strongly associated with measures of cardiac structure and function among males and females at age 25 years. Earlier puberty is unlikely to have a major impact on pre-clinical cardiovascular risk in early adulthood.

## Data Availability

The study website contains details of all the data that is available through a fully searchable data dictionary and variable search tool: http://www.bristol.ac.uk/alspac/researchers/our-data/

## Acknowledgements

We are extremely grateful to all the families who took part in this study, the midwives for their help in recruiting them, and the whole ALSPAC team, which includes interviewers, computer and laboratory technicians, clerical workers, research scientists, volunteers, managers, receptionists and nurses.

