## Supplement for "Puberty Timing and Markers of Cardiovascular Structure and Function at 25 Years: A Prospective Cohort Study"

**Online-Only Supplement**

**eMethods 1:** Description of confounding variables.

**eTable 1:** Variables used in multivariable multiple imputation models.

**eTable 2:** Likelihood ratio test examining linearity of association between age at peak height velocity and cardiac structure and function outcomes by sex.

**eTable 3:** Distributions of imputed characteristics in the imputation datasets and in observed data (i.e. without imputation) for CIMT in males and females.

**eTable 4:** Distributions of imputed characteristics in the imputation datasets and in observed data (i.e. without imputation) for LVMI in males and females.
**eTable 5:** Distributions of imputed characteristics in the imputation datasets and in observed data (i.e. without imputation) for RWT in males and females.
 **eTable 6:** Distributions of imputed characteristics in the imputation datasets and in observed data (i.e. without imputation) for PWV in males and females.
 **eTable 7:** Distributions of imputed characteristics in the imputation datasets and in observed data (i.e. without imputation) for SBP in males and females.
**eTable 8:** Adjusted associations of aPHV with measures of cardiac structure and function among participants with complete-case data on exposure, outcome and covariates.

**eMethods 1**

**Description of confounding variables**

Maternal age was reported in the mother’s antenatal questionnaires. Gestational age at birth was estimated from clinical records. Household social class was measured as the highest of the mother’s or her partner’s occupational social class using data on job title and details of occupation collected about the mother and her partner from the mother’s questionnaire at 32 weeks gestation. Social class was derived using the standard occupational classification (SOC) codes developed by the United Kingdom Office of Population Census and Surveys and classified as I professional, II managerial and technical, IIINM non-manual, IIIM manual, and IV&V part skilled occupations and unskilled occupations.

A questionnaire at 32 weeks gestation asked mothers to report on educational attainment, which was categorized as below O-Level (Ordinary Level; exams taken in different subjects usually at age 15-16 at the completion of legally required school attendance, equivalent to today’s UK General Certificate of Secondary Education), O-Level only, A-Level (Advanced-Level; exams taken in different subjects usually at age 18), or university degree or above.

Breastfeeding information was collected via questionnaires administered at 4 weeks, 6 months and 15 months. Parity was defined as the number of previous pregnancies that had resulted in a live or stillborn infant collected at 18 weeks gestation. Birthweight was extracted from medical records. Maternal height and weight data were self-reported from a questionnaire administered at 12 weeks gestation; these were used to calculate maternal BMI. Marital status was obtained from antenatal questionnaires and classified as never married, married, and widowed, divorced, or separated. Smoking in the first trimester of pregnancy was self-reported by mothers at 18 weeks gestation; responses to smoking any tobacco (cigarettes, cigars, pipes, or other) were grouped as follows: no smoking, <10 per day, 10-19 per day or greater than 19 per day, and were re-categorised as a dichotomous variable (smoking: yes/no).

Height and fat mass of offspring was measured at clinics at age 9 years. Standing height was measured to the last complete mm using the Harpenden Stadiometer. Fat mass (in kg, less head) was derived from whole body DXA scans performed using a GE Lunar Prodigy (Madison, WI, USA) narrow fan beam densitometer.

**eTable 1 Variables used in multivariable multiple imputation models**

| **Variable** | **Type of variable** | **Model used to**  **predict missing**  **data in this variable** | **How variable**  **entered when used**  **to predict missing**  **in other variables** |
| --- | --- | --- | --- |
| aPHV | Continuous | n/a, complete case was used | Continuous |
| CIMT at age 17 | Continuous | Linear regression | Continuous |
| CIMT at age 25 | Continuous | Linear regression | Continuous |
| LVMI at age 17 | Continuous | Linear regression | Continuous |
| LVMI at age 25 | Continuous | Linear regression | Continuous |
| RWT at age 17 | Continuous | Linear regression | Continuous |
| RWT at age 25 | Continuous | Linear regression | Continuous |
| PWV at age 17 | Continuous | Linear regression | Continuous |
| PWV at age 25 | Continuous | Linear regression | Continuous |
| SBP at age 17 | Continuous | Linear regression | Continuous |
| SBP at age 25 | Continuous | Linear regression | Continuous |
| Maternal age | Continuous | Linear regression | Continuous |
| Gestational age | Ordered categorical  (10 categories) | Ordinal logistic  regression | 9 indicator  variables |
| Household social class | Categorical  (4 categories) | Multinomial logistic  regression | 3 indicator  variables |
| Maternal education | Categorical  (4 categories) | Multinomial logistic  regression | 3 indicator  variables |
| Paternal education | Categorical  (4 categories) | Multinomial logistic  regression | 3 indicator  variables |
| Breastfeeding | Categorical  (3 levels) | Multinomial  logistic regression | 2 indicator variables |
| Birthweight | Continuous | Linear regression | Continuous |
| Maternal BMI | Continuous | Linear regression | Continuous |
| Maternal smoking | Categorical  (2 levels) | Multinomial logistic  regression | 1 indicator variable |
| Height at age 9 | Continuous | Linear regression | Continuous |
| Fat mass at age 9 | Continuous | Linear regression | Continuous |
| Parity | Categorical  (3 levels) | Multinomial  logistic regression | 2 indicator variables |
| Marital status | Categorical  (3 levels) | Multinomial  logistic regression | 2 indicator variables |
| Abbreviations: aPHV, age at peak height velocity; CIMT, carotid intima-media thickness; LVMI, left ventricular mass index; RWT, relative wall thickness; PWV, pulse wave velocity; SBP, systolic blood pressure; BMI, body mass index. | | | |

**eTable 2 Likelihood ratio test examining linearity of association between age at peak height velocity and cardiac structure and function outcomes by sex**

|  | **Males** | **Females** |
| --- | --- | --- |
|  | **P value comparing models** | **P value comparing models** |
| CIMT | 0.82 | 0.18 |
| LVMI | 0.39 | 0.61 |
| RWT | 0.15 | 0.52 |
| PWV | 0.33 | 0.28 |
| SBP | 0.59 | 0.73 |

P-value from likelihood ratio test comparing fit of models regressing CIMT, LVMI, RWT, PWV and SBP on fourths of aPHV (treated as a continuous exposure) to models regressing CIMT, LVMI, RWT, PWV and SBP on fourths of aPHV (treated as a categorical exposure).

P>0.05 indicates the more parsimonious model (aPHV treated as a continuous exposure) is a better fit, suggesting linearity of associations of aPHV and each of the outcomes.

**eTable 3: Distributions of imputed characteristics in the imputation datasets (N=4,339) and in observed data (N=1199) (i.e. without imputation) for CIMT in males and females**

| **Imputed variable** | **% imputed data** | **Distribution**  **% for categorical variables**  **Mean (SE) for continuous variables** | | **% imputed data** | **Distribution**  **% for categorical variables**  **Mean (SE) for continuous variables** | |
| --- | --- | --- | --- | --- | --- | --- |
|  | **Males** | | | **Females** | | |
|  |  | **Imputed dataset** | **Observed data (without imputation)** |  | **Imputed dataset** | **Observed data (without imputation)** |
| ***Household social class*** |  |  |  |  |  |  |
| Professional | 10.1 | 19.94 | 25.32 | 10.2 | 17.29 | 18.90 |
| Managerial/technical |  | 47.63 | 47.47 |  | 45.29 | 47.59 |
| Non-manual |  | 21.43 | 18.57 |  | 23.83 | 24.28 |
| Manual/part skills/unskilled |  | 11.00 | 8.65 |  | 13.59 | 9.24 |
| ***Maternal education*** |  |  |  |  |  |  |
| Less than O level | 6.7 | 17.01 | 12.03 | 7.0 | 17.94 | 12.14 |
| O level |  | 32.08 | 29.75 |  | 34.92 | 37.24 |
| A level |  | 30.26 | 32.91 |  | 27.77 | 28.00 |
| Degree or above |  | 20.65 | 25.32 |  | 19.37 | 22.62 |
| ***Mother’s partner’s education*** |  |  |  |  |  |  |
| Less than O level | 8.6 | 21.64 | 17.72 | 9.2 | 26.49 | 21.38 |
| O level |  | 22.21 | 17.51 |  | 21.01 | 21.38 |
| A level |  | 28.99 | 29.75 |  | 28.74 | 28.55 |
| Degree or above |  | 27.16 | 35.02 |  | 23.76 | 28.69 |
| ***Breastfeeding until 3 months*** |  |  |  |  |  |  |
| Exclusively | 10.5 | 34.57 | 37.34 | 11.7 | 38.16 | 40.14 |
| Non-exclusively |  | 53.53 | 52.11 |  | 46.83 | 45.66 |
| Never |  | 11.90 | 10.55 |  | 15.01 | 14.21 |
| ***First-born child*** | 6.6 | 49.92 | 49.37 | 8.0 | 48.37 | 48.14 |
| ***Maternal marital status*** |  |  |  |  |  |  |
| Never married | 5.9 | 11.99 | 8.44 | 6.1 | 13.33 | 8.00 |
| Married |  | 83.34 | 87.13 |  | 81.96 | 87.45 |
| Widowed/divorced/separated |  | 4.67 | 4.43 |  | 4.71 | 4.55 |
| ***Maternal smoking status*** |  |  |  |  |  |  |
| No | 5.9 | 84.62 | 87.55 | 6.3 | 84.84 | 87.59 |
| Yes |  | 15.38 | 12.45 |  | 15.16 | 12.41 |
| Maternal age at delivery (years) | 4.6 | 29.66 (0.10) | 30.15 (0.19) | 4.8 | 29.36 (0.09) | 29.88 (0.15) |
| Gestational age (weeks) | 4.6 | 39.32 (0.04) | 39.59 (0.07) | 4.8 | 39.51 (0.03) | 39.67 (0.05) |
| Birthweight (kg) | 5.5 | 3.49 (0.01) | 3.53 (0.02) | 6.0 | 3.38 (0.01) | 3.37 (0.01) |
| Maternal BMI (kg/m^2^) | 11.5 | 22.91 (0.08) | 22.88 (0.16) | 12.7 | 22.74 (0.07) | 22.72 (0.13) |
| Height at age 9 (cm) | 4.9 | 139.93 (0.16) | 139.92 (0.27) | 5.7 | 139.02 (0.15) | 139.16 (0.22) |
| Fat mass at age 9 (kg) | 8.3 | 33.92 (0.15) | 33.66 (0.29) | 9.1 | 34.18 (0.15) | 34.04 (0.25) |
| Age at peak height velocity (years) | n/a, complete case was used | 13.54 (0.02) | 13.49 (0.04) | n/a, complete case was used | 11.73 (0.01) | 11.75 (0.03) |
| CIMT at age 25 (mm) | 66.0 | 0.46 (0.001) | 0.46 (0.002) | 56.4 | 0.45 (0.001) | 0.45 (0.001) |
| Abbreviations: BMI, body mass index; CIMT, carotid intima-media thickness. | | | | | | |

**eTable 4: Distributions of imputed characteristics in the imputation datasets (N=2752) and in observed data (N=1197) (i.e. without imputation) for LVMI in males and females**

| **Imputed variable** | **% imputed data** | **Distribution**  **% for categorical variables**  **Mean (SE) for continuous variables** | | **% imputed data** | **Distribution**  **% for categorical variables**  **Mean (SE) for continuous variables** | |
| --- | --- | --- | --- | --- | --- | --- |
|  | **Males** | | | **Females** | | |
|  |  | **Imputed dataset** | **Observed data (without imputation)** |  | **Imputed dataset** | **Observed data (without imputation)** |
| ***Household social class*** |  |  |  |  |  |  |
| Professional | 9.0 | 21.56 | 23.58 | 9.1 | 18.75 | 20.08 |
| Managerial/technical |  | 47.51 | 50.53 |  | 44.58 | 48.06 |
| Non-manual |  | 20.21 | 18.11 |  | 25.13 | 23.82 |
| Manual/part skills/unskilled |  | 10.69 | 7.79 |  | 11.52 | 8.03 |
| ***Maternal education*** |  |  |  |  |  |  |
| Less than O level | 6.0 | 16.35 | 10.53 | 6.0 | 17.00 | 11.77 |
| O level |  | 31.49 | 29.89 |  | 33.88 | 34.76 |
| A level |  | 30.25 | 34.53 |  | 28.33 | 30.06 |
| Degree or above |  | 21.89 | 25.05 |  | 20.78 | 23.41 |
| ***Mother’s partner’s education*** |  |  |  |  |  |  |
| Less than O level | 7.6 | 19.31 | 15.79 | 8.0 | 23.77 | 20.23 |
| O level |  | 23.14 | 19.58 |  | 20.90 | 21.61 |
| A level |  | 28.77 | 30.74 |  | 28.53 | 27.98 |
| Degree or above |  | 28.75 | 33.89 |  | 26.77 | 30.19 |
| ***Breastfeeding until 3 months*** |  |  |  |  |  |  |
| Exclusively | 8.9 | 34.37 | 36.42 | 10.3 | 39.17 | 41.00 |
| Non-exclusively |  | 54.07 | 54.32 |  | 46.84 | 45.29 |
| Never |  | 11.55 | 9.26 |  | 13.98 | 13.71 |
| ***First-born child*** | 6.1 | 51.83 | 48.84 | 7.1 | 48.45 | 48.34 |
| ***Maternal marital status*** |  |  |  |  |  |  |
| Never married | 5.3 | 11.79 | 8.21 | 5.6 | 13.13 | 9.00 |
| Married |  | 83.22 | 87.16 |  | 82.50 | 86.57 |
| Widowed/divorced/separated |  | 4.98 | 4.63 |  | 4.35 | 4.43 |
| ***Maternal smoking status*** |  |  |  |  |  |  |
| No | 5.2 | 86.77 | 88.84 | 5.7 | 85.15 | 87.40 |
| Yes |  | 13.22 | 11.16 |  | 14.84 | 12.60 |
| Maternal age at delivery (years) | 4.2 | 29.74 (0.13) | 30.36 (0.20) | 4.3 | 29.53 (0.11) | 29.90 (0.15) |
| Gestational age (weeks) | 4.2 | 39.39 (0.05) | 39.52 (0.07) | 4.3 | 39.54 (0.04) | 39.65 (0.05) |
| Birthweight (kg) | 5.0 | 3.48 (0.01) | 3.51 (0.02) | 5.4 | 3.38 (0.01) | 3.36 (0.01) |
| Maternal BMI (kg/m^2^) | 10.4 | 22.88 (0.11) | 22.66 (0.16) | 13.0 | 22.71 (0.09) | 22.52 (0.12) |
| Height at age 9 (cm) | 4.9 | 139.77 (0.15) | 139.74 (0.27) | 5.0 | 138.94 (0.14) | 138.97 (0.22) |
| Fat mass at age 9 (kg) | 8.7 | 33.68 (0.19) | 33.08 (0.27) | 8.0 | 34.05 (0.18) | 33.76 (0.24) |
| Age at peak height velocity (years) | n/a, complete case was used | 13.51 (0.02) | 13.55 (0.04) | n/a, complete case was used | 11.75 (0.02) | 11.77 (0.03) |
| LVMI at age 25 (g/m^2.7^) | 44.4 | 32.68 (0.28) | 32.55 (0.31) | 33.2 | 29.32 (0.18) | 28.86 (0.23) |
| Abbreviations: BMI, body mass index; LVMI, left ventricular mass index. | | | | | | |

**eTable 5: Distributions of imputed characteristics in the imputation datasets (N=2776) and in observed data (N=1203) (i.e. without imputation) for RWT in males and females**

| **Imputed variable** | **% imputed data** | **Distribution**  **% for categorical variables**  **Mean (SE) for continuous variables** | | **% imputed data** | **Distribution**  **% for categorical variables**  **Mean (SE) for continuous variables** | |
| --- | --- | --- | --- | --- | --- | --- |
|  | **Males** | | | **Females** | | |
|  |  | **Imputed dataset** | **Observed data (without imputation)** |  | **Imputed dataset** | **Observed data (without imputation)** |
| ***Household social class*** |  |  |  |  |  |  |
| Professional | 8.9 | 21.37 | 23.43 | 9.1 | 18.53 | 20.00 |
| Managerial/technical |  | 48.03 | 50.42 |  | 44.52 | 48.14 |
| Non-manual |  | 20.21 | 18.20 |  | 25.41 | 23.86 |
| Manual/part skills/unskilled |  | 10.37 | 7.95 |  | 11.51 | 8.00 |
| ***Maternal education*** |  |  |  |  |  |  |
| Less than O level | 5.9 | 16.32 | 10.46 | 5.9 | 17.06 | 11.86 |
| O level |  | 31.43 | 30.13 |  | 34.02 | 34.76 |
| A level |  | 30.39 | 34.52 |  | 28.22 | 30.07 |
| Degree or above |  | 21.84 | 24.90 |  | 20.69 | 23.31 |
| ***Mother’s partner’s education*** |  |  |  |  |  |  |
| Less than O level | 7.7 | 18.64 | 15.89 | 8.0 | 23.14 | 20.13 |
| O level |  | 23.00 | 19.67 |  | 20.94 | 21.52 |
| A level |  | 29.00 | 30.96 |  | 28.64 | 28.14 |
| Degree or above |  | 29.36 | 33.68 |  | 27.26 | 30.21 |
| ***Breastfeeding until 3 months*** |  |  |  |  |  |  |
| Exclusively | 9.0 | 34.61 | 36.19 | 10.2 | 38.80 | 40.97 |
| Non-exclusively |  | 53.91 | 54.60 |  | 47.13 | 45.38 |
| Never |  | 11.46 | 9.21 |  | 14.06 | 13.66 |
| ***First-born child*** | 6.2 | 52.13 | 49.16 | 7.0 | 48.72 | 48.41 |
| ***Maternal marital status*** |  |  |  |  |  |  |
| Never married | 5.2 | 11.82 | 8.16 | 5.5 | 13.17 | 9.10 |
| Married |  | 83.22 | 87.24 |  | 82.31 | 86.48 |
| Widowed/divorced/separated |  | 4.94 | 4.60 |  | 4.51 | 4.41 |
| ***Maternal smoking status*** |  |  |  |  |  |  |
| No | 5.2 | 86.57 | 88.91 | 5.7 | 85.09 | 87.45 |
| Yes |  | 13.43 | 11.09 |  | 14.91 | 12.55 |
| Maternal age at delivery (years) | 4.1 | 29.75 (0.13) | 30.32 (0.20) | 4.2 | 29.52 (0.11) | 29.91 (0.15) |
| Gestational age (weeks) | 4.1 | 39.38 (0.05) | 39.52 (0.07) | 4.2 | 39.54 (0.04) | 39.64 (0.05) |
| Birthweight (kg) | 5.0 | 3.48 (0.01) | 3.51 (0.02) | 5.4 | 3.37 (0.01) | 3.37 (0.01) |
| Maternal BMI (kg/m^2^) | 10.4 | 22.89 (0.11) | 22.67 (0.16) | 12.9 | 22.72 (0.09) | 22.53 (0.12) |
| Height at age 9 (cm) | 4.8 | 139.75 (0.17) | 139.73 (0.27) | 5.1 | 138.94 (0.16) | 138.98 (0.22) |
| Fat mass at age 9 (kg) | 8.7 | 33.70 (0.19) | 33.08 (0.27) | 8.0 | 34.05 (0.18) | 33.76 (0.24) |
| Age at peak height velocity (years) | n/a, complete case was used | 13.51 (0.02) | 13.55 (0.04) | n/a, complete case was used | 11.76 (0.02) | 11.77 (0.03) |
| RWT at age 25 | 44.5 | 0.36 (0.001) | 0.36 (0.002) | 33.5 | 0.35 (0.001) | 0.35 (0.002) |
| Abbreviations: BMI, body mass index; RWT, relative wall thickness. | | | | | | |

**eTable 6: Distributions of imputed characteristics in the imputation datasets (N=3964) and in observed data (N=1394) (i.e. without imputation) for PWV in males and females**

| **Imputed variable** | **% imputed data** | **Distribution**  **% for categorical variables**  **Mean (SE) for continuous variables** | | **% imputed data** | **Distribution**  **% for categorical variables**  **Mean (SE) for continuous variables** | |
| --- | --- | --- | --- | --- | --- | --- |
|  | **Males** | | | **Females** | | |
|  |  | **Imputed dataset** | **Observed data (without imputation)** |  | **Imputed dataset** | **Observed data (without imputation)** |
| ***Household social class*** |  |  |  |  |  |  |
| Professional | 9.9 | 20.78 | 23.59 | 9.7 | 16.90 | 19.34 |
| Managerial/technical |  | 46.65 | 49.36 |  | 46.25 | 48.40 |
| Non-manual |  | 21.55 | 18.51 |  | 23.93 | 23.25 |
| Manual/part skills/unskilled |  | 10.99 | 8.53 |  | 12.90 | 9.02 |
| ***Maternal education*** |  |  |  |  |  |  |
| Less than O level | 6.7 | 16.59 | 10.89 | 6.6 | 17.60 | 12.46 |
| O level |  | 32.25 | 31.22 |  | 34.84 | 36.18 |
| A level |  | 30.78 | 32.67 |  | 28.29 | 28.94 |
| Degree or above |  | 20.37 | 25.23 |  | 19.24 | 22.42 |
| ***Mother’s partner’s education*** |  |  |  |  |  |  |
| Less than O level | 8.5 | 21.25 | 15.61 | 8.5 | 25.70 | 21.35 |
| O level |  | 22.38 | 20.15 |  | 21.04 | 20.64 |
| A level |  | 28.88 | 31.40 |  | 28.91 | 29.54 |
| Degree or above |  | 27.46 | 32.85 |  | 24.33 | 28.47 |
| ***Breastfeeding until 3 months*** |  |  |  |  |  |  |
| Exclusively | 10.3 | 33.90 | 35.39 | 11.4 | 39.22 | 40.33 |
| Non-exclusively |  | 53.68 | 53.72 |  | 46.68 | 45.91 |
| Never |  | 12.41 | 10.89 |  | 14.09 | 13.76 |
| ***First-born child*** | 6.8 | 49.83 | 50.82 | 7.6 | 48.42 | 48.64 |
| ***Maternal marital status*** |  |  |  |  |  |  |
| Never married | 6.0 | 11.54 | 8.53 | 5.9 | 12.82 | 9.37 |
| Married |  | 83.56 | 87.11 |  | 82.37 | 86.00 |
| Widowed/divorced/separated |  | 4.89 | 4.36 |  | 4.80 | 4.63 |
| ***Maternal smoking status*** |  |  |  |  |  |  |
| No | 5.8 | 84.83 | 88.57 | 6.2 | 85.15 | 87.78 |
| Yes |  | 15.16 | 11.43 |  | 14.84 | 12.22 |
| Maternal age at delivery (years) | 4.7 | 29.68 (0.11) | 30.05 (0.18) | 4.5 | 29.45 (0.09) | 29.81 (0.14) |
| Gestational age (weeks) | 4.7 | 39.31 (0.04) | 39.53 (0.06) | 4.5 | 39.52 (0.03) | 39.60 (0.05) |
| Birthweight (kg) | 5.6 | 3.48 (0.01) | 3.51 (0.02) | 5.7 | 3.38 (0.01) | 3.37 (0.01) |
| Maternal BMI (kg/m^2^) | 11.1 | 22.83 (0.09) | 22.80 (0.15) | 12.6 | 22.66 (0.07) | 22.65 (0.11) |
| Height at age 9 (cm) | 4.6 | 139.87 (0.14) | 139.85 (0.25) | 5.5 | 139.07 (0.13) | 139.17 (0.21) |
| Fat mass at age 9 (kg) | 8.1 | 33.72 (0.15) | 33.64 (0.26) | 8.9 | 34.09 (0.15) | 34.08 (0.23) |
| Age at peak height velocity (years) | n/a, complete case was used | 13.53 (0.02) | 13.50 (0.03) | n/a, complete case was used | 11.74 (0.01) | 11.76 (0.02) |
| PWV at age 25 (m/s) | 56.3 | 6.63 (0.04) | 6.72 (0.05) | 44.3 | 6.10 (0.02) | 6.12 (0.03) |
| Abbreviations: BMI, body mass index; PWV, pulse wave velocity. | | | | | | |

**eTable 7: Distributions of imputed characteristics in the imputation datasets (N=4571) and in observed data (2193) (i.e. without imputation) for SBP in males and females**

| **Imputed variable** | **% imputed data** | **Distribution**  **% for categorical variables**  **Mean (SE) for continuous variables** | | **% imputed data** | **Distribution**  **% for categorical variables**  **Mean (SE) for continuous variables** | |
| --- | --- | --- | --- | --- | --- | --- |
|  | **Males** | | | **Females** | | |
|  |  | **Imputed dataset** | **Observed data (without imputation)** |  | **Imputed dataset** | **Observed data (without imputation)** |
| ***Household social class*** |  |  |  |  |  |  |
| Professional | 10.5 | 19.85 | 23.34 | 10.3 | 17.25 | 19.83 |
| Managerial/technical |  | 47.14 | 48.82 |  | 45.40 | 46.32 |
| Non-manual |  | 22.70 | 19.84 |  | 24.18 | 23.58 |
| Manual/part skills/unskilled |  | 10.29 | 8.00 |  | 13.15 | 10.26 |
| ***Maternal education*** |  |  |  |  |  |  |
| Less than O level | 6.7 | 16.93 | 12.51 | 7.1 | 17.71 | 12.10 |
| O level |  | 33.02 | 32.02 |  | 35.31 | 35.45 |
| A level |  | 29.75 | 31.79 |  | 28.27 | 30.63 |
| Degree or above |  | 20.28 | 23.68 |  | 18.68 | 21.82 |
| ***Mother’s partner’s education*** |  |  |  |  |  |  |
| Less than O level | 8.7 | 21.58 | 17.82 | 9.2 | 26.74 | 22.21 |
| O level |  | 22.55 | 19.95 |  | 20.91 | 20.29 |
| A level |  | 28.91 | 30.21 |  | 28.49 | 29.63 |
| Degree or above |  | 26.94 | 32.02 |  | 23.83 | 27.87 |
| ***Breastfeeding until 3 months*** |  |  |  |  |  |  |
| Exclusively | 10.6 | 34.63 | 37.88 | 11.6 | 38.13 | 40.28 |
| Non-exclusively |  | 53.09 | 52.20 |  | 47.11 | 45.94 |
| Never |  | 12.27 | 9.92 |  | 14.74 | 13.78 |
| ***First-born child*** | 6.6 | 50.31 | 50.85 | 7.7 | 48.45 | 48.62 |
| ***Maternal marital status*** |  |  |  |  |  |  |
| Never married | 6.0 | 12.20 | 10.03 | 6.2 | 13.58 | 10.49 |
| Married |  | 83.24 | 86.02 |  | 81.97 | 85.91 |
| Widowed/divorced/separated |  | 4.54 | 3.95 |  | 4.43 | 3.60 |
| ***Maternal smoking status*** |  |  |  |  |  |  |
| No | 5.7 | 84.49 | 87.49 | 6.2 | 85.13 | 88.28 |
| Yes |  | 15.51 | 12.51 |  | 14.87 | 11.72 |
| Maternal age at delivery (years) | 4.5 | 29.62 (0.10) | 30.02 (0.14) | 4.7 | 29.32 (0.09) | 29.61 (0.11) |
| Gestational age (weeks) | 4.5 | 39.32 (0.04) | 39.48 (0.05) | 4.7 | 39.51 (0.03) | 39.63 (0.04) |
| Birthweight (kg) | 5.4 | 3.49 (0.01) | 3.50 (0.01) | 5.9 | 3.38 (0.01) | 3.39 (0.01) |
| Maternal BMI (kg/m^2^) | 11.4 | 22.93 (0.08) | 22.90 (0.12) | 12.7 | 22.71 (0.07) | 22.62 (0.09) |
| Height at age 9 (cm) | 5.2 | 139.98 (0.13) | 140.06 (0.20) | 5.8 | 139.05 (0.13) | 139.03 (0.17) |
| Fat mass at age 9 (kg) | 8.3 | 34.04 (0.15) | 33.94 (0.22) | 8.9 | 34.20 (0.15) | 34.03 (0.19) |
| Age at peak height velocity (years) | n/a, complete case was used | 13.53 (0.02) | 13.48 (0.03) | n/a, complete case was used | 11.74 (0.01) | 11.75 (0.02) |
| SBP at age 25 (mmHg) | 38.1 | 122.80 (0.27) | 123.10 (0.36) | 24.8 | 111.66 (0.21) | 111.76 (0.27) |
| Abbreviations: BMI, body mass index; SBP, systolic blood pressure. | | | | | | |

**eTable 8: Adjusted associations of aPHV with measures of cardiac structure and function among participants with complete-case data on exposure, outcome and covariates**

|  | **Males** | **Females** |
| --- | --- | --- |
| **Outcome** | **Estimate (95% CI)^a,b^** | **Estimate (95% CI)^a,b^** |
| **CIMT** | 0.006 (0.001, 0.01) | -0.002 (-0.007, 0.001) |
| **LVMI** | -0.18 (-0.87, 0.50) | -1.53 (-0.90, -2.16) |
| **RWT** | -0.002 (-0.008, 0.003) | -0.006 (-0.0001, -0.01) |
| **PWV** | 0.02 (-0.09, 0.14) | 0.04 (-0.05, 0.13) |
| **SBP** | 0.25 (-0.62, 1.14) | 0.01 (-0.72, 0.76) |
| ^a^Association of a one-year older aPHV with each outcome.  ^b^Adjusted for maternal age, gestational age, household social class, maternal education, mother’s partner’s education, breastfeeding of baby until three months, parity, birthweight, maternal body mass index, maternal marital status, maternal smoking status during first 3 months of pregnancy, and height and fat mass of offspring at age 9.  Abbreviations: aPHV, age at peak height velocity; CI, confidence interval; CIMT, carotid intima-media thickness; LVMI, left ventricular mass index; RWT, relative wall thickness; PWV, pulse wave velocity; SBP, systolic blood pressure. | | |
